# Emergency department visits and boarding for pediatric patients with suicidality before and during the COVID-19 pandemic

**DOI:** 10.1101/2023.05.08.23289659

**Authors:** Amy R. Zipursky, Karen L. Olson, Louisa Bode, Alon Geva, James Jones, Kenneth D. Mandl, Andrew McMurry

## Abstract

**Objective:** To quantify the increase in pediatric patients presenting to the emergency department with suicidality before and during the COVID-19 pandemic, and the subsequent impact on emergency department length of stay and boarding.

**Methods:** This retrospective cohort study from June 1, 2016, to October 31, 2022, identified patients presenting to the emergency department with suicidality using ICD-10 codes. Number of emergency department encounters for suicidality, demographic characteristics of patients with suicidality, and emergency department length of stay were compared before and during the COVID-19 pandemic. Unobserved components models were used to describe monthly counts of emergency department encounters for suicidality.

**Results:** There were 179,736 patient encounters to the emergency department during the study period, 6,168 (3.4%) for suicidality. There were, on average, more encounters for suicidality each month during the COVID-19 pandemic than before the COVID-19 pandemic. A time series unobserved components model demonstrated an initial drop in encounters for suicidality in April and May of 2020, followed by an increase starting in July 2020. The average length of stay for patients that boarded in the emergency department with a diagnosis of suicidality was 37.4 hours longer during the COVID-19 pandemic compared to before the COVID-19 pandemic.

**Conclusions:** The number of encounters for suicidality among pediatric patients and the emergency department length of stay for psychiatry boarders has increased during the COVID-19 pandemic. There is a need for acute care mental health services and solutions to emergency department capacity issues.

## Introduction

Suicide is the second leading cause of death for 10- to 14-year-olds and the third leading cause of death for 15- to 19-year-olds [1]. Since the start of the COVID-19 pandemic [2–8], an increase in suicidal ideation and attempt among pediatric patients has been well-documented, resulting in an increase in emergency department (ED) visits [7,9] and straining EDs in the United States [10].

Patients visiting the ED for mental health issues are twice as likely to be hospitalized compared to other patients [11]. Because there is a shortage of inpatient psychiatric beds and limited outpatient mental health services, patients presenting to the ED for mental health reasons often board in the ED for long periods of time [10,12]. This problem has been exacerbated during the COVID-19 pandemic, with increased waiting times for pediatric psychiatric beds [13]. A survey of hospitals in the United States and Canada found that 75% of those that responded noted an increase in boarding duration and 84% an increase in boarding frequency during the COVID-19 pandemic [12]. Patients boarding in the ED often do not have access to the psychiatric services they need for optimal care [12]. The urgent need for solutions to ED boarding and mental health capacity issues has been recognized [10,14]. Many studies to date have focused on the first year of the pandemic, and have not examined boarding and capacity issues. We sought to quantify the increase in pediatric patients presenting to the ED with suicidality during the COVID-19 pandemic, and the subsequent impact on ED length of stay (LOS) and boarding.

## Methods

This is a retrospective cohort study using the electronic health records (EHR) of patients who presented to the ED at Boston Children’s Hospital (BCH), a pediatric academic medical center, between June 1, 2016 and October 31, 2022. Patients between 6 and 21 years old were included and age was recorded as an integer [15]. Patients with multiple ED visits were counted once for each visit and could be included in different age groups over the course of the study.

The study received exempt status from Institutional Review Board at BCH.

<u>Suicidality</u> is defined using International Classification of Diseases, 10th Revision (ICD-10) codes selected from those used in prior studies [16–23]. We included codes for suicidal ideation (R45.851), suicide attempt (T14.91*), and codes for intentional self-harm. A comprehensive list is in S1 Table.

<u>ED LOS</u> is defined as the time between ED check-in and check-out. Prolonged LOS was defined as more than 24 hours.

<u>Psychiatry boarders</u> are patients who needed a higher level of psychiatric care before discharge from the ED. They were identified by the existence of a standard psychiatry boarder form in the EHR that was completed by a provider.

<u>COVID-19 pandemic periods</u> were defined as pre-COVID-19 (June 1, 2016 to February 29, 2020) and COVID-19 (March 1, 2020 to October 31, 2022) [24].

<u>Race and ethnicity</u> were self-reported by patients or their parents or guardians at intake. They could select one or more racial categories including other and indicate their ethnicity as Hispanic or Latino if applicable. For this study, race and ethnicity were coded as mutually exclusive categories: Hispanic/Latino (any race group), and Non-Hispanic/Latino groups: Asian, Black/African American, Multiracial, Other Race, White, and a final category for Unknown.

<u>Seasons</u> were defined as winter (January, February, March), spring (April, May, June), summer (July, August, September), and fall (October, November, December).

### Analysis

The number of ED encounters with ICD-10 codes for suicidal ideation, suicide attempt, and intentional self-harm pre-COVID-19 and during COVID-19 were compared. We also compared the demographic characteristics of patients during these two time periods. ED LOS for encounters with suicidality diagnoses were compared for those with and without psychiatry boarders during the two time periods. Analyses used Chi-square and analysis of variance (ANOVA) with post-hoc Tukey tests as appropriate. Least squares means (LSM) and 95% confidence limits (CL) or differences between LSMs are reported for ANOVA results. Analyses were performed using SAS 9.4, Copyright (c) 2020 by SAS Institute Inc., Cary, NC.

Unobserved components models (UCM) were used to describe monthly counts of ED encounters for suicidality. Time series can be described in terms of their components such as trend, seasonality, and cycles, and can include regression effects [25–28]. Our initial model included components for trend and seasonality. Regression variables were added to assess the impact of COVID-19 in 2020, with April 2020 as the initial month of impact. Statistical change point analysis was used to evaluate other notable trend changes. Analyses were performed using the UCM Procedure in SAS 9.4. P values less than 0.05 were considered significant.

## Results

There were 179,736 ED encounters during the study period and 6,168 (3.4%) had an ICD-10 code for suicidality. ANOVA revealed main effects for the COVID-19 pandemic period and season (p<0.001 for each) and no significant interaction. On average, there were more suicidality encounters each month during the COVID-19 time period (LSM 101.2, CL 94.7, 107.7) than during the pre-COVID-19 time period (LSM 66.7, CL 61.3, 72.2). Compared to summer months, on average there were 19.3 (CL 3.9, 34.6) more suicidality encounters during spring, 30.7 (CL 15.0, 46.3) more during fall, and 33.3 (CL 17.5, 49.1) more during winter months.

Among all ED encounters, we examined how many suicidality encounters versus all others were characterized by a prolonged LOS or a psychiatry boarder form in the EHR. More suicidality encounters (2,905, 47.1%) had a prolonged ED LOS than encounters without that diagnosis (2,627, 1.5%, p<0.001). More suicidality encounters had a completed boarder form (4,692, 76.1%) than other ED encounters (4,730, 2.7%, p<0.001).

The demographic characteristics of patients with a suicidality diagnosis during ED encounters in the pre-COVID-19 and COVID-19 time periods are shown in Table 1. The majority of encounters (84.6%) were with patients 12-18 years old. Differences between the COVID-19 pandemic periods were not significant (p=0.655). The majority of patients were female, and they comprised a greater percentage of patients during the COVID-19 time period (71.5%) than during the pre-COVID-19 time period (65.3%). Race and ethnicity was reported as White, non-Hispanic for the majority of patients (3,243, 52.6%) during suicidality encounters, followed by Hispanic/Latino (914, 14.8%) and Black/African American, non-Hispanic (763, 12.4%). The distribution of categories varied somewhat between the time periods (p<0.001). Post-hoc analyses indicated that only one category, Unknown, was statistically different.

**Table 1.** Characteristics of patients with a suicidality diagnosis during ED encounters before and during the COVID-19 pandemic.

| Variable | Values | Pre-COVID-19<br>N 2,986 |  | COVID-19<br>N 3,182 |  | p-value |
| --- | --- | --- | --- | --- | --- | --- |
|  |  | Number | Percent | Number | Percent |  |
| **Age (years)** | 6-11 | 361 | 12.1 | 361 | 11.3 | 0.655 |
|  | 12-18 | 2,515 | 84.2 | 2,705 | 85.0 |  |
|  | 19-21 | 110 | 3.7 | 116 | 3.6 |  |
| **Sex** | Female | 1,949 | 65.3 | 2,276 | 71.5 | <0.001 |
|  | Male | 1,037 | 34.7 | 906 | 28.5 |  |
| **Race and ethnicity <sup>a</sup>** | Asian, non-Hispanic | 125 | 4.2 | 108 | 3.4 | <0.001 |
|  | Black/African American, non-Hispanic | 399 | 13.4 | 364 | 11.4 |  |
|  | Hispanic/Latino | 453 | 15.2 | 461 | 14.5 |  |
|  | Multiracial, non-Hispanic | 49 | 1.6 | 59 | 1.9 |  |
|  | Other race, non-Hispanic | 212 | 7.1 | 175 | 5.5 |  |
|  | White, non-Hispanic | 1,565 | 52.4 | 1,678 | 52.7 |  |
|  | Unknown <sup>b</sup> | 183 | 6.1 | 337 | 10.6 |  |

<sup>a</sup> Categories are mutually exclusive.
<sup>b</sup> Significant difference between pre-COVID-19 and COVID-19 time periods in post-hoc analyses.

Suicide attempt was coded for 322 encounters (5.2%) and these encounters could also include codes for intentional self-harm (118), ideation (68), or both (51). Intentional self-harm codes, without an attempt code, described 941 (15.3%) encounters, 428 with an ideation code. Ideation without attempt or intentional self-harm codes described the majority (4,905, 79.5%) of encounters. There were no differences by time period (p=0.121).

The most frequent intentional self-harm diagnosis codes were poisoning by 4-Aminophenol derivatives, observed in 234 (3.8%) of all encounters with suicidality, poisoning by unspecified drugs, medications, and biological substances (N 220, 3.6%), poisoning by propionic acid derivatives (N 161, 2.6%), self-harm by unspecified sharp object (N 146, 2.4%), and poisoning by selective serotonin reuptake inhibitors (N 142, 2.3%). For the 1,100 encounters with an intentional self-harm code, we compared the top five codes in three conditions: pre-COVID-19 vs. COVID-19 time period, encounters with and without a diagnosis for suicide attempt, and encounters with and without a diagnosis for suicide ideation (without attempt). The top five for each comparison were mostly the same as those included in the overall list above, sometimes with different rank orders. And in two cases, another code replaced one of the top five. Therefore, seven codes were compared in three conditions and we used a Bonferroni-adjusted alpha value of 0.002. Six differences, listed in Table 2, were observed. Full details regarding the self-harm codes are in S2-5 Tables.

**Table 2.** Differences observed for three comparisons of the most frequent intentional self-harm diagnosis codes.

| Comparison | Code description | Result |
| --- | --- | --- |
| **Pre-COVID-19 vs. COVID-19 time period** | Intentional self-harm by unspecified sharp object | Higher during COVID-19 (N 110 of 568, 19.4%, rank 2) than before (N 36 of 542, 6.6%, rank 6). |
|  | Intentional self-harm by other specified means | Lower during COVID-19 (N 42 of 568, 7.4%, rank 6) than before (N 76 of 542), 14.0%, rank 3). |
| **Diagnosis for suicide attempt vs. no diagnosis for attempt** | Intentional self-harm by unspecified sharp object | Lower for encounters with a diagnosis code for suicide attempt (N 8 of 169, 4.7%, rank 10) than encounters without a diagnosis for attempt (N 138 of 941, 14.7%, rank 3). |
|  | Asphyxiation due to hanging | Higher for encounters with diagnosis code for suicide attempt (N 13 of 169, 7.7%, rank 5) than encounters without a diagnosis for attempt (N 6 of 941, 0.6%, rank 27). |
| **Diagnosis for suicide ideation, without attempt vs. no diagnosis for ideation or attempt** | Poisoning by propionic acid derivatives, intentional self-harm | Higher for encounters with a diagnosis code for suicide ideation (without attempt) (N 74 of 428, 17.3%, rank 2) than encounters without a diagnosis for ideation (N 52 of 513, 10.1%, rank 6). |
|  | Intentional self-harm by other specified means | Lower for encounters with a diagnosis code for suicide ideation (without attempt) (N 28 of 428, 6.5%, rank 6) than encounters without a diagnosis for ideation (N 81 of 513, 15.8%, rank 3). |

The effect of psychiatry boarder status and COVID-19 pandemic period on ED LOS is shown in Table 3. ANOVA revealed significant main effects for boarder status and time period, and a significant interaction (p<0.001 for all). Encounters without psychiatry boarders were not prolonged and post-hoc analyses showed that ED LOS did not differ between the pre-COVID-19 and COVID-19 time periods. However, encounters with boarders were, on average, prolonged and those during the COVID-19 pandemic were 37.4 hours longer than before the COVID-19 pandemic. Also, before the COVID-19 pandemic, suicidality encounters where the patient was boarding were 22.2 hours longer than encounters without boarding. During the COVID-19 pandemic, this difference was 61.8 hours.

**Table 3.**
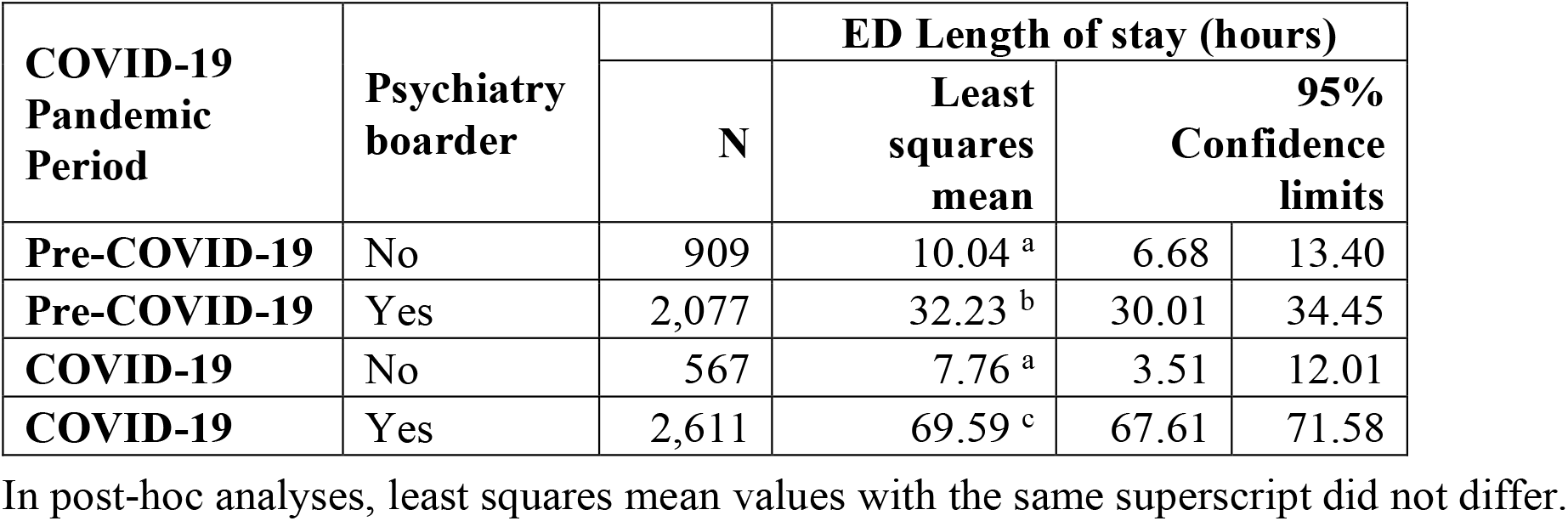
The effect of psychiatry boarder status and COVID-19 pandemic period on ED LOS for patients with a suicidality diagnosis.

| COVID-19<br>Pandemic<br>Period | Psychiatry<br>boarder | N | ED Length of stay (hours) |  |  |
| --- | --- | --- | --- | --- | --- |
|  |  |  | Least<br>squares<br>mean | 95%<br>Confidence<br>limits |  |
| Pre-COVID-19 | No | 909 | 10.04<sup>a</sup> | 6.68 | 13.40 |
| Pre-COVID-19 | Yes | 2,077 | 32.23<sup>b</sup> | 30.01 | 34.45 |
| COVID-19 | No | 567 | 7.76<sup>a</sup> | 3.51 | 12.01 |
| COVID-19 | Yes | 2,611 | 69.59<sup>c</sup> | 67.61 | 71.58 |

In post-hoc analyses, least squares mean values with the same superscript did not differ.

A time series of the number of ED suicidality encounters is shown in Fig 1. Seasonality is apparent, with lowest points in summer months, except during 2020. The UCM analysis for suicidality encounters included a trend component modeled as a random walk with drift, deterministic seasonality, and two intervention variables. A smoothed plot of the seasonality component is shown in Fig 2. The estimate for level at its final state was 72.4 encounters (standard error (se) 12.1, p<0.001), while the slope was close to zero (0.2, se 0.6, p=0.730). The trend was effectively a random walk with little or no drift. Two intervention variables described the impact of COVID-19. A temporary change lasting two months (April-May 2020) had an effect size of -33.0 encounters (se 9.3, p<0.001). A level change beginning in July 2020 had a sustained effect size of 31.1 (se 10.6, p=0.003). Fig 3 shows the smoothed sum of the trend and regression effects (April-May 2020 drop, increase since July 2020) by isolating these effects from the complete series. A smoothed plot of the sum of all the components and regression effects, minus the irregular term, is shown in Fig 4.

**Fig 1.**
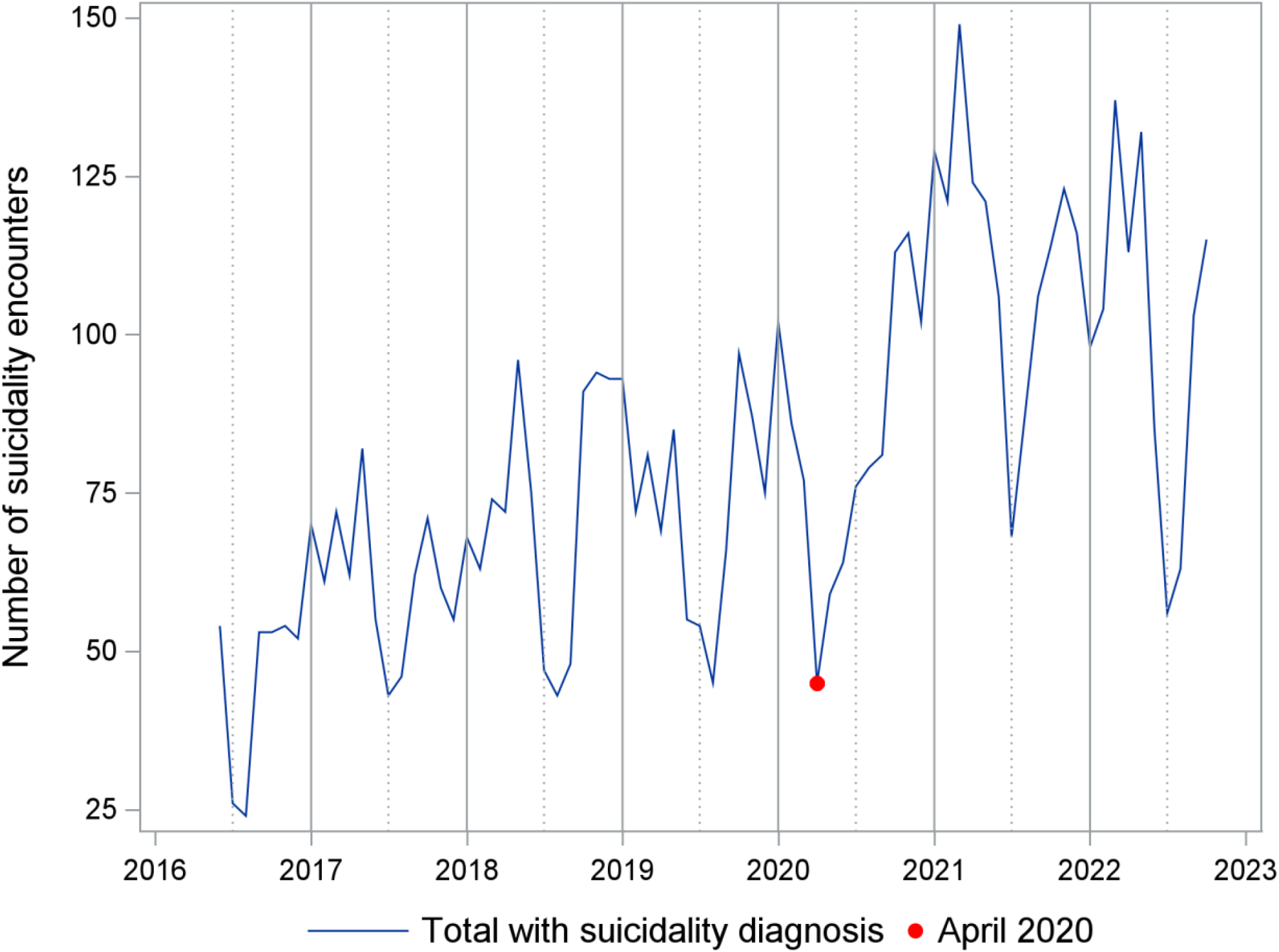
Time series for number of emergency department encounters with suicidality. Number of encounters per month between June 1, 2016 and October 31, 2022.

**Fig 2.**
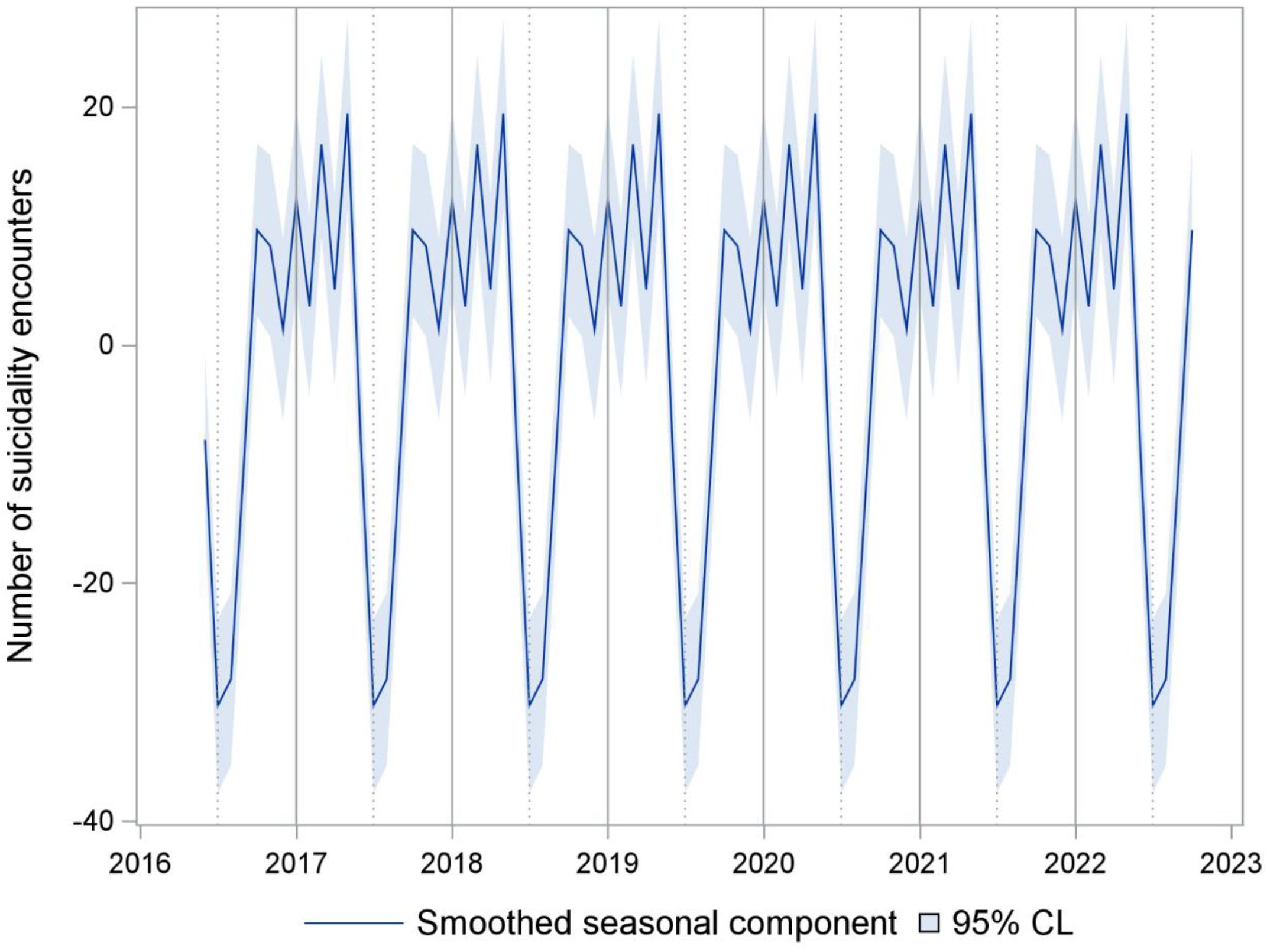
Smoothed seasonal component for emergency department encounters with suicidality. CL= Confidence limits. Seasonal component of a time series unobserved components model.

**Fig 3.**
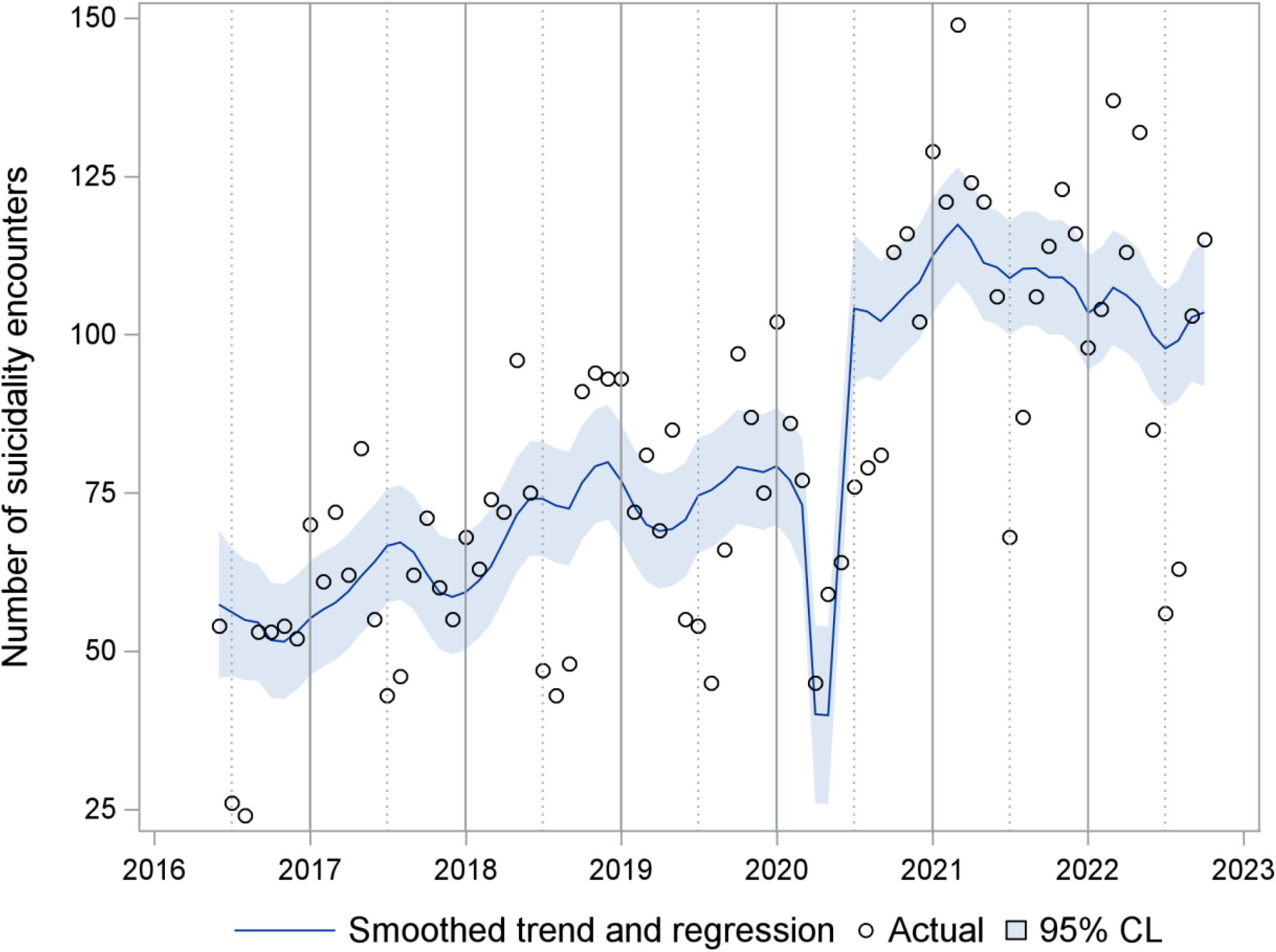
Smoothed trend and regression effects (April-May 2020 drop, July 2020 level shift). CL= Confidence limits. Sum of effects from a time series unobserved components model for emergency department encounters with suicidality.

**Fig 4.**
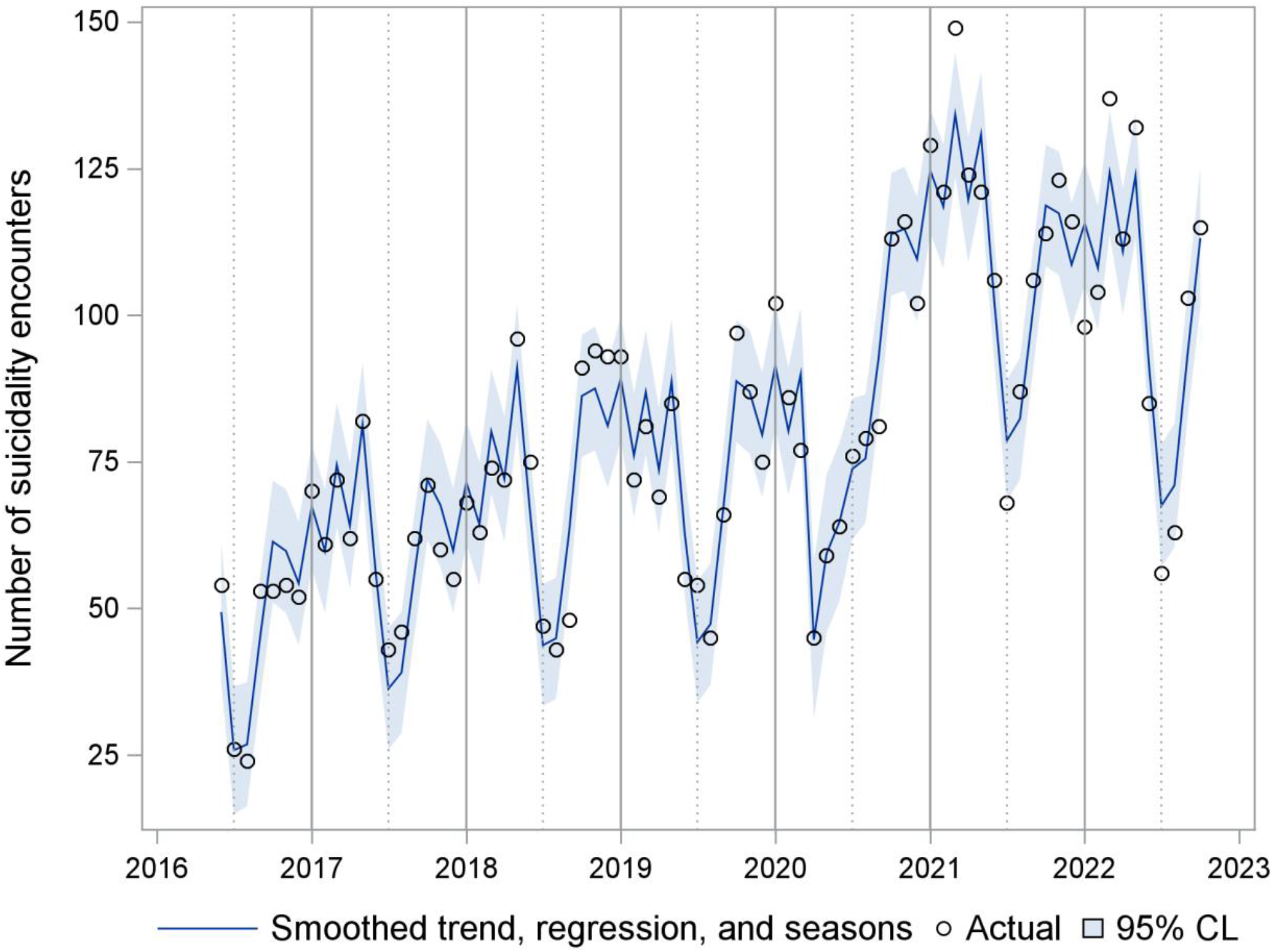
UCM sum of the smoothed trend, regression, and season effects. CL= Confidence limits. Sum of all components except the irregular term of a time series unobserved components model for emergency department encounters with suicidality. Regression effects are a drop in April and May 2020 and a level shift beginning in July 2020.

## Discussion

After an initial drop at the beginning of the COVID-19 pandemic in April and May 2020, the number of ED encounters for suicidality has increased during the COVID-19 pandemic.

Previous studies have shown a decrease in ED visits overall and ED visits for psychiatric issues early in the COVID-19 pandemic [6,29,30]. Individuals may have accessed care less frequently early in the COVID-19 pandemic due to fear of contracting COVID-19. The subsequent increase we observed in ED visits for suicidality is consistent with previous literature. Existing literature, mainly focusing on data up to 2021 [2,4–7,13], found an increase in pediatric suicidality during the COVID-19 pandemic as well as increased ED visits [2–7,13] and hospitalizations [31]. We likely saw an increase in presentations for suicidality as a result of persistent stressors related to COVID-19 on youth mental health as well as delayed presentations from early in the pandemic. Our results also describe an increase in the number and duration of ED boarding for those presenting to the ED with suicidality. Previous literature has described an increase in mental health boarding in general through 2021 and has included ED and inpatient unit stays or other temporary location stays in the definition of boarding [12,13]. This study specifically highlights capacity issues related to the ED and patients presenting with suicidality.

Furthermore, with data through October 2022, our study demonstrates the sustained impact of COVID-19 on pediatric suicidality.

This study has some limitations. First, while we chose diagnostic codes used in the published literature to identify patients with suicidality [16–23], these ICD-10 codes are nonetheless intended for patient billing. Second, though our time series demonstrates a compelling trend, other secular changes during the COVID-19 pandemic may not be accounted for. Pediatric primary care was limited at points during COVID-19 pandemic due to factors such as access to personal protective equipment and ability to isolate infectious patients [32]. This may have led patients to present to the ED rather than accessing the primary care settings that they would have prior to the COVID-19 pandemic. Finally, our study was in a large, well-resourced children’s hospital in an urban setting. While studies conducted in other settings have found comparable results <u>[2,4–7,29]</u>, these findings may not generalize to all settings.

Overall, we demonstrate an increase in patients with suicidality that presented to the ED during the COVID-19 pandemic and the capacity issues facing the ED. Future studies should focus on expanding this work to multiple medical centers. This work supports the need for an increase in acute care mental health services and solutions to ED capacity issues.

## Data Availability

All data produced in the present study are available upon reasonable request to the authors.

## Acknowledgements

None

## Supporting Information

**S1 Table. ICD-10 search list for suicide attempt, suicide ideation, and intentional self-harm**

**S2 Table. Number of ED encounters for self-harm ICD-10 codes observed in the data**

**S3 Table. Number of ED encounters with self-harm codes by COVID-19 pandemic period**

**S4 Table. Number of ED encounters with self-harm codes by suicide attempt (yes/no)**

**S5 Table. Number of ED encounters with self-harm codes by suicide ideation without attempt (yes/no)**

